# Using surgical wrapping material for the fabrication of respirator masks

**DOI:** 10.1101/2020.04.10.20060632

**Authors:** Johanna H Meijer, Joric Oude Vrielink

## Abstract

Given the current shortage of respirator masks and the resulting lack of personal protective equipment for use by clinical staff, we examined bottom-up solutions that would allow hospitals to fabricate respirator masks that: (i) meet requirements in terms of filtering capacities, (ii) are easy to produce rapidly and locally, and (iii) can be constructed using materials commonly available in hospitals worldwide. We found that Halyard H300 material used for wrapping of surgical instruments and routinely available in hospitals, met these criteria. Specifically, three layers of material achieved a filter efficiency of 94%, 99%, and 100% for 0.3 μm, 0.5 μm, and 3.0 μm particles, respectively; importantly, these values are close to the efficiency provided by FFP2 and N95 masks. After re-sterilization up to 5 times, the filter’s efficiency remains sufficiently high for use as an FFP1 respirator mask. Finally, using only one layer of the material satisfies the criteria for use as a ‘surgical mask’. This material can therefore be used to help protect hospital staff and other healthcare professionals who require access to suitable masks but lack commercially available solutions.

## Introduction

The SARS-CoV-2 pandemic, which began in late 2019 and rapidly spread throughout the world in early 2020, has led to a severe shortage of basic personal protective equipment for people working in high-risk occupations, including hospitals and extended care homes. Because SARS-CoV-2 can be transmitted via aerosolized droplets [1], the demand for high-quality, well tested respirator masks has increased dramatically. In an attempt to help provide access to personal protective equipment against SARS-CoV-2, particularly in locations currently experiencing a shortage of masks [2], we investigated whether sterilization wrapping material is suitability for use in the production of FFP2 masks. We chose to use this material because of its filter properties, and its current availability in most hospitals, and the near-global sales market. The wrapping material Halyard Quickcheck H300 (manufactured by Owens & Minor) is commonly used to wrap surgical instruments for sterilization with steam or ethylene oxide gas; after sterilization, the material provides a sterile barrier against pathogens. Thus, the filter properties of this wrapping material are similar to the materials used in respirator masks. Moreover, the surgical wrapping material is composed of polypropylene, a material commonly used in air filters. Although sterilization wrapping has previously been suggested as an alternative material for the local production of respirator masks [3][4][5], it has not been tested.

A series of industrial standards are currently used to classify the filter efficiency of respirators. The European Norm (EN) 149 standard [6] classifies respirators using FFP (Filtering Facemask against Particles) values, while World Health Organisation (WHO) guidelines require that healthcare workers—particularly those working under aerosol-generating conditions—wear a mask that provides at least FFP2 protection or equivalent [7], with FFP2 corresponding to a filter efficiency of at least 94%. Providing a filter efficiency of at least 95%, similar standards are used for N95 and NK95 masks, corresponding to regulation NIOSH 42 CFR 84 in the US and norm GB2626-2006 in China, respectively. For respirator masks, the material’s efficiency at filtering particles 0.3 μm in size is considered the most relevant, as particles of this size are the extremely difficult to filter out [8].

In addition to the above-mentioned EN 149 norm, several commercially available respirators are also classified using the EN 14683 standard [9]; the equivalent classification in the US is the ASTM F2100 standard. This standard is specific to masks used for medical purposes, primarily to protect patients from contamination, for example during surgery. In light of the SARS-CoV-2 pandemic, the WHO now recommends the use of surgical masks by patients who are potentially infected with the virus and hospital staff who work with these patients [10]. As an alternative to surgical masks, FFP1 respirators have also been suggested; however, although FFP1 respirator masks provide better particle protection than surgical masks, they are less splash-resistant than surgical masks [12].

Here, we investigated whether Halyard wrapping material can be used to create masks in order to help protect healthcare workers under various conditions. We used standardized equipment to measure the transmission of various particle sizes, and we also measured splash resistance. Finally, we used standard sterilization procedures in order to investigate whether the material can be re-used while still providing sufficient protection. Moreover, we developed a prototype for a complete, functional respirator mask, with the aim of facilitating its rapid on-site fabrication. Importantly, we also identified additional materials that allow for the production of high quantities of respirator masks using basic workshop instruments and found that the masks can be produced in at the relatively rapid rate of under 10 minutes per mask.

## Methods

The filter properties of the surgical isolation and wrapping material Halyard Quickcheck H300 (Owens & Minor, Inc.) was tested using a Solair 3100 particle counter (Lighthouse Worldwide Solutions Benelux B.V.) as shown in Figure 1. The flow rate was set at 1.0 CFM (cubic feet per minute)cfm, which is well above (4x) normal breathing, and the filter efficiency of the material for 0.3 μm, 0.5 μm, and 3.0 μm particles was measured. The filter efficiency (*FE*) for a given particle size was determined using the particles measured after filtering (*P_f_*) with a background measurement performed without the filter *(P_BG_*), using the following equation: *FE = (P_BG_ - P_f_)/P_BG_*.

**Figure 1.**
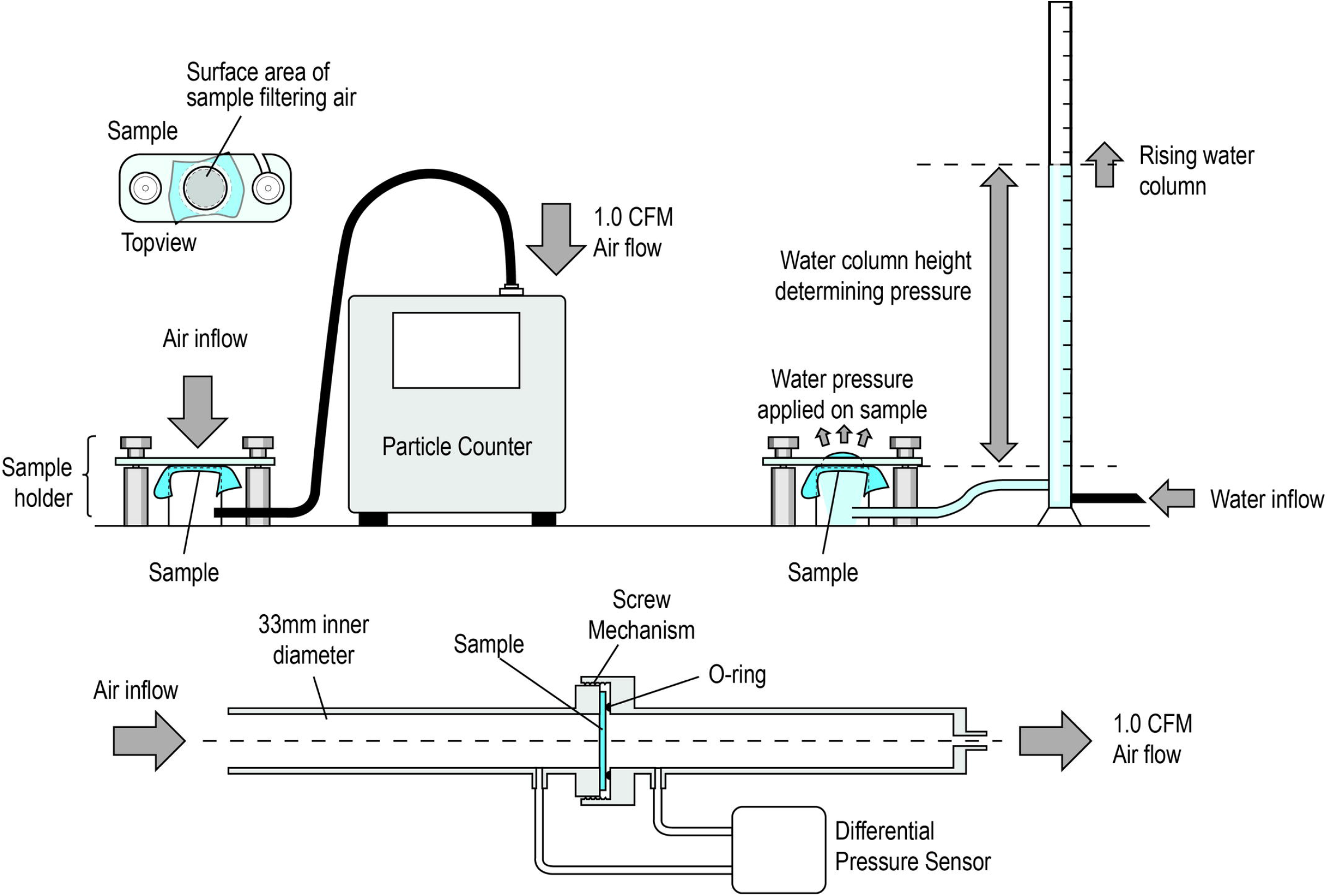
Schematic diagrams depicting the tests used to measure filter efficiency (top left), splash resistance (top right), and breathability of the materials examined in this study.

In addition, the pressure differential was measured over the sample to provide a measure of breathing resistance. The surface of the material through which the air passes is a circle with 33 mm diameter; this surface area was used to normalize the measured pressure to Pa/cm^2^. For comparison, we also measured the differential pressure of commercially available FFP2 respirators and surgical masks.

Splash resistance was measured using a water column pressure test (Figure 1). In brief, the water column is filled gradually, increasing the pressure at the sample, and the height of water in the column withheld by the material sample is defined as the sample’s splash resistance.

We tested 1, 2, and 3 layers of Halyard Quick Check H300 material and performed each measurement four times. All tests were then repeated following each subsequent round of steam sterilization (5 min at 121 degrees Celsius under 2.0 atm of pressure). In addition, these tests were repeated with the material reversed and after the mask was worn for 20 min. Finally, three commercially available masks were tested as a control and for comparison with the custom-made masks: (1) Surgical Mask, Henan Gore Medical Instruments Co. Ltd (2) Disposable FFP2 mask, (3) 3820 FFP2 NR D, 3M.

This study did not involve any human subjects, and therefore was exempt from ethical approval. The data from the Particle Counter was downloaded and stored into a .xslx worksheet. The data of the differential pressure test and the water column test was manually inserted in separate .xlsx files. The data analyses was performed using Microsoft Excel, including the independent t-tests required for statistical analysis.

## Results

Figure 2 shows the results obtained using various configurations of wrapping material. Three layers of non-sterilized wrapping material provided mean (±SD) filter efficiency values of 93.84±0.37%, 99.45±0.08%, and 99.99±0.01% for 0.3 μm, 0.5 μm, and 3.0 μm particles, respectively, satisfying the criterion for an FFP2 respirator mask (summary results are provided in Table 1, and the complete results are provided in the Supplemental Data). Two layers resulted in 87.68±0.43%, 98.28±0.10%, and 99.98±0.00%, respectively, was satisfies the criteria for an FFP1 respirator mask. Importantly, the triple-layer material’s efficiency at filtering 0.5 μm and 3.0 μm particles remained above the FFP2 standards even after five rounds of sterilization cycles; however, the filter efficiency for 0.3 μm particles decreased below the requirements for an FFP2 mask, but was still sufficient for use an as FFP1 mask.

**Figure 2.**
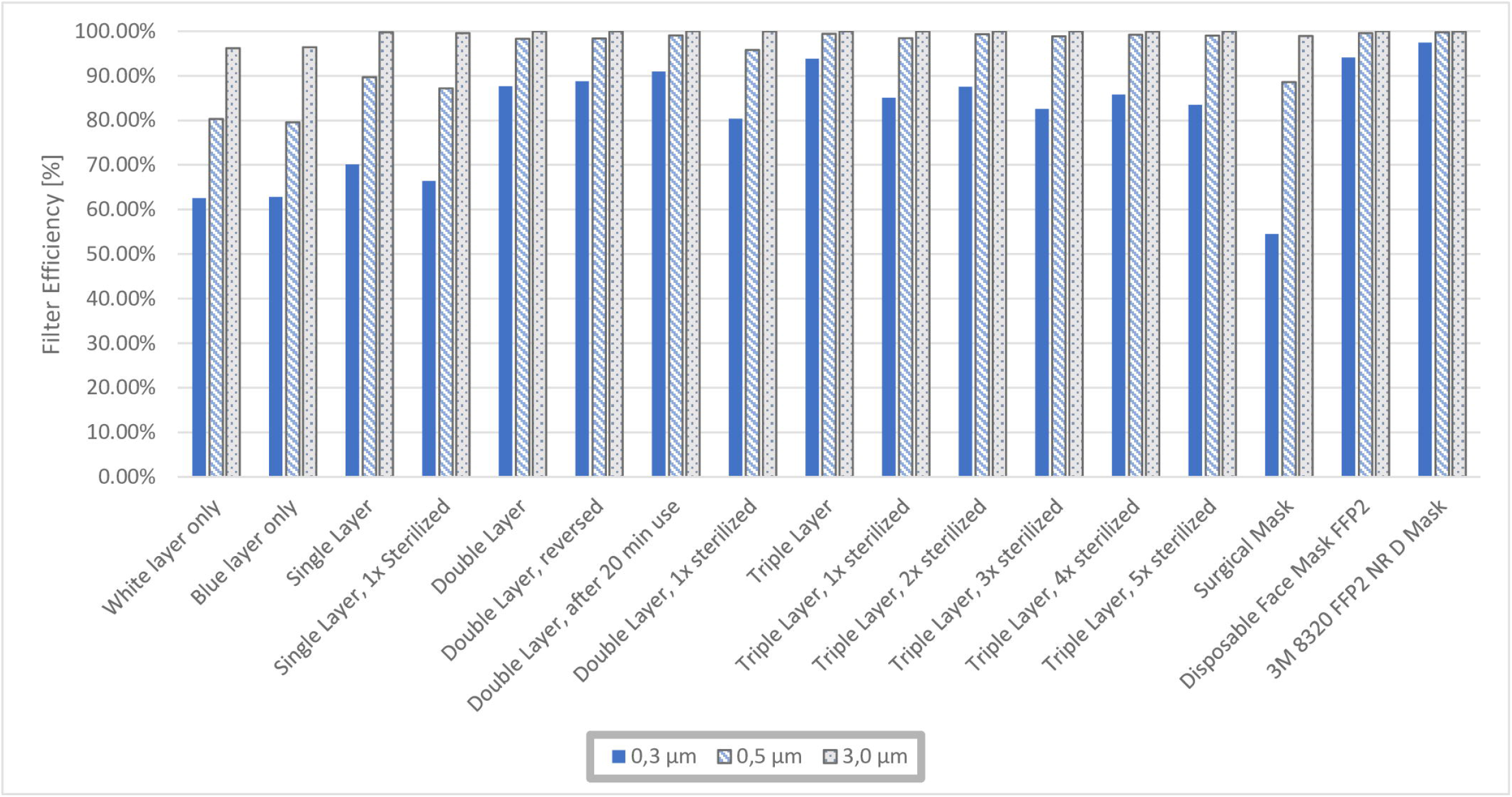
Summary of the filter efficiency of the indicated materials, including five rounds of sterilization at 121 degrees Celsius for the triple-layer material. For comparison, we also included a surgical mask and two types of FFP2 mask.

**Table 1.**
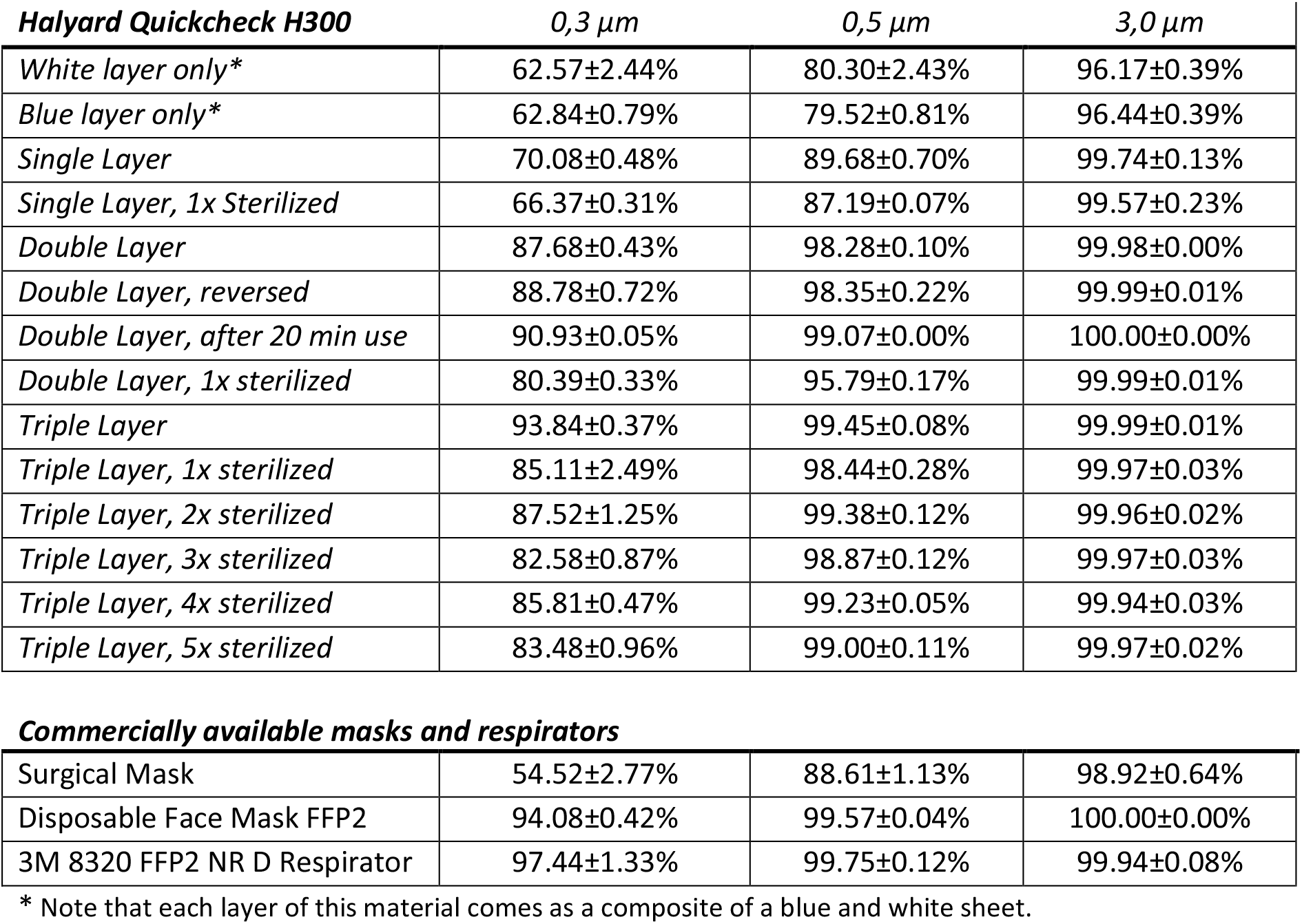
Summary of the filter efficiency of the Quickcheck H300 material and commercially available masks, for the indicated particle sizes.

| **Halyard Quickcheck H300** | **0,3 μm** | **0,5 μm** | **3,0 μm** |
| --- | --- | --- | --- |
| White layer only* | 62.57±2.44% | 80.30±2.43% | 96.17±0.39% |
| Blue layer only* | 62.84±0.79% | 79.52±0.81% | 96.44±0.39% |
| Single Layer | 70.08±0.48% | 89.68±0.70% | 99.74±0.13% |
| Single Layer, 1x Sterilized | 66.37±0.31% | 87.19±0.07% | 99.57±0.23% |
| Double Layer | 87.68±0.43% | 98.28±0.10% | 99.98±0.00% |
| Double Layer, reversed | 88.78±0.72% | 98.35±0.22% | 99.99±0.01% |
| Double Layer, after 20 min use | 90.93±0.05% | 99.07±0.00% | 100.00±0.00% |
| Double Layer, 1x sterilized | 80.39±0.33% | 95.79±0.17% | 99.99±0.01% |
| Triple Layer | 93.84±0.37% | 99.45±0.08% | 99.99±0.01% |
| Triple Layer, 1x sterilized | 85.11±2.49% | 98.44±0.28% | 99.97±0.03% |
| Triple Layer, 2x sterilized | 87.52±1.25% | 99.38±0.12% | 99.96±0.02% |
| Triple Layer, 3x sterilized | 82.58±0.87% | 98.87±0.12% | 99.97±0.03% |
| Triple Layer, 4x sterilized | 85.81±0.47% | 99.23±0.05% | 99.94±0.03% |
| Triple Layer, 5x sterilized | 83.48±0.96% | 99.00±0.11% | 99.97±0.02% |
| **Commercially available masks and respirators** |  |  |  |
| Surgical Mask | 54.52±2.77% | 88.61±1.13% | 98.92±0.64% |
| Disposable Face Mask FFP2 | 94.08±0.42% | 99.57±0.04% | 100.00±0.00% |
| 3M 8320 FFP2 NR D Respirator | 97.44±1.33% | 99.75±0.12% | 99.94±0.08% |

\* Note that each layer of this material comes as a composite of a blue and white sheet.

Table 1 shows the filtering efficiencies for the wrapping material. Based on the tests with the double layer material it is shown that the filter efficiency did not significantly change when reversing the material or after wearing the respirator mask for 20 min (p = 0.057 for 0.3*μm* particle size). After sterilization of the wrapping material the filter efficiency at the 0.3 μm decreased significantly after 1 round of sterilization for single, double and triple layers of materials (p =0.019, p = 0.004 and p = 0.000, respectively). The filter efficiency for 3.0μm particles in triple layer material significantly changed after 4 sterilization cycles (p = 0.049).

As shown in Figure 3 (supplementary data in S2 Data), the pressure differential across the wrapping material, which provides an approximation of the material’s breathability, increases with each additional layer). No significant difference in pressure differential was observed after the sterilization (p_1_ = 0.099, p_2_ = 0.540, p_3_ = 0.327, p_4_ = 0.961, p_5_ = 0.577 for the 1-5 sterilization cycles compared to unsterilized material, respectively).

**Figure 3.**
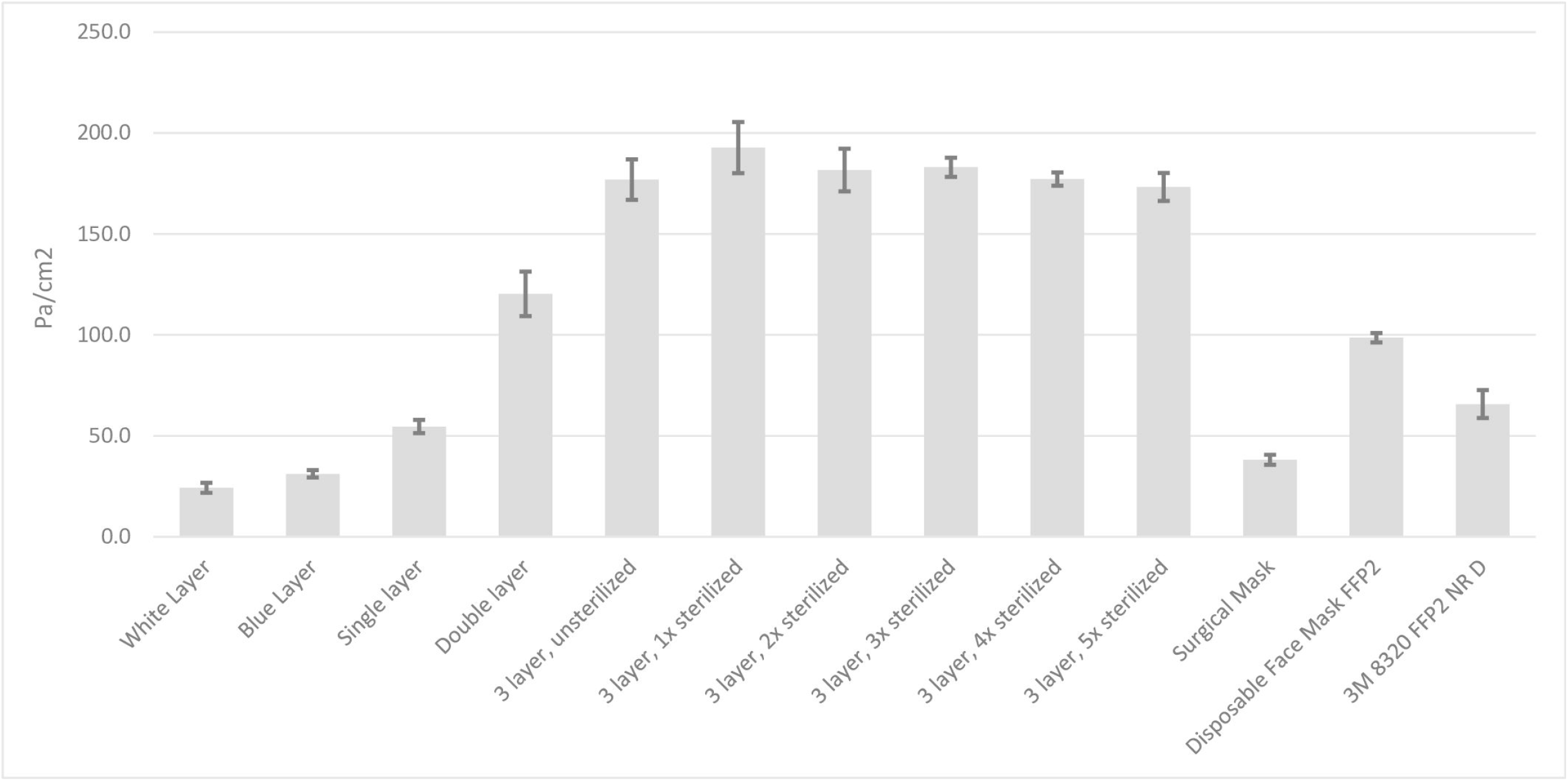
The differential pressure over the different layers of materials, including after different rounds of sterilization. As a comparison the differential pressure is compared to commercially available surgical masks and disposable FFP2 respirators.

Figure 4 summarizes the results of the water column test, which provides an approximate measure of the materials’ splash resistance (Supplementary data in S3 Data). With two and three layers of wrapping material, the mean column pressure was 92 and 105 cm H_2_O, respectively; similar results were obtained when the material was reversed.

**Figure 4.**
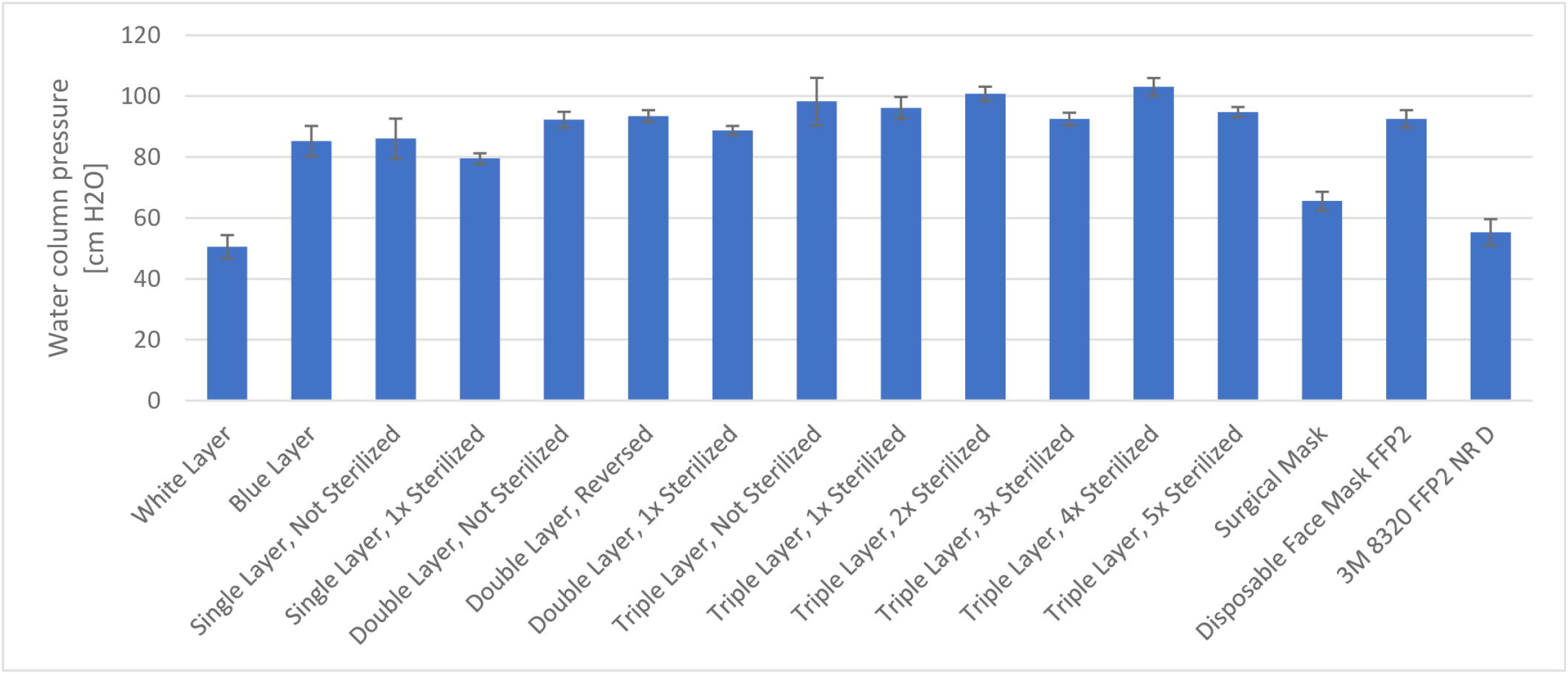
Summary of the results of the water column test.

A single layer of non-sterilized wrapping material provided similar or better results compared to a surgical mask in terms of filter efficiency (p = 0.001), and splash resistance (p = 0.002). On the other hand, the breathability of the surgical mask was better than the wrapping material (p = 0.000, one-tailed independent t-test).

As a comparison Halyard Quickcheck H500, which is used for wrapping heavier instrumentation, was tested (supplementary S4 data). The material shows improved splash resistance and filter efficiencies, also for 0.3 μm, however, the material has considerable higher breathing resistance than the H300 material.

## Discussion

Here, we report that the Halyard Quickcheck H300 surgical wrapping material, which is commonly available in most hospitals for use in sterilizing surgical instruments, provides high filtration efficiency for all three particle sizes measured. Specifically, three layers of the material filtered approximately 94% of 0.3 μm particles, which is similar to the requirements established for N95 (95%) and FFP2 (94%) respirator masks [13][14]. In addition, the three-layer provided a filter efficiency of 99.5% and 100% for 0.5 and 3.0 μm particles, respectively. Moreover, we found that the wrapping material provides high splash resistance; although this is not a requirement for respirator masks, this property helps protect against transmission due to coughing and sneezing. After several sterilization rounds, the masks were suitable for use as FFP1 respirators.

In addition, we found that a single layer of wrapping material is suitable for use as a high-quality alternative for splash resistant surgical mask. Of note, our testing method differed from the method recommended by EN 14683; specifically, we did not test bacterial pathogens. The filter efficiency of surgical masks is determined specifically based on the ability to filter the *Staphylococcus aureus* bacterium, which has a diameter of 0.5-1.5 μm [12] [15]. Although we measured particles, rather than bacteria, we found that even a single layer of H300 provides higher filter efficiencies and splash resistance.

The precise particle size that is most critical in the transmission of SARS-CoV-2 is currently under debate and subject to change as new information becomes available. The current prevailing view is that somewhat larger-diameter aerosols play a major role in the virus’ transmission but that also small aerosols or particles can carry the virus over long distances [16]. These aerosols can reach the ACE-2 receptor proteins abundantly expressed in the upper respiratory tract [17][18], and coughing and sneezing are believed to be one of the major routes of transmission [19]. In this respect, the high splash resistance of H300 material seems particularly important, and although this property is not an official requirement of N95 and FFP masks, we believe that this will support its protective function.

The breathability of the H300 material decreased with each layer added. We found that three layers of material have high breathing resistance, above the maximum standard for surgical masks, which is provided as a measure per surface area. Notably, breathability for N95 or FFP2 respirator masks is not specified for the material but for the entire mask, making it dependent on the design and surface area.

We constructed a complete respiratory mask in order to facilitate in-house production. We identified a basic set of conventional materials (aluminum, neoprene rubber, and elastic material) with suitable specifications (see Figure 5), recognizing that many other designs will also work [3][4][5]. Most hospitals—even in rural areas—have a basic workshop. The only key requirement for our design is a machine to cut aluminum and elastic strips out of larger sheets. The model that we propose is relatively easy to make, and we intentionally designed the mask so that the wearer can breathe through the entire surface of the mask, increasing breathability and thereby comfort.

**Figure 5.**
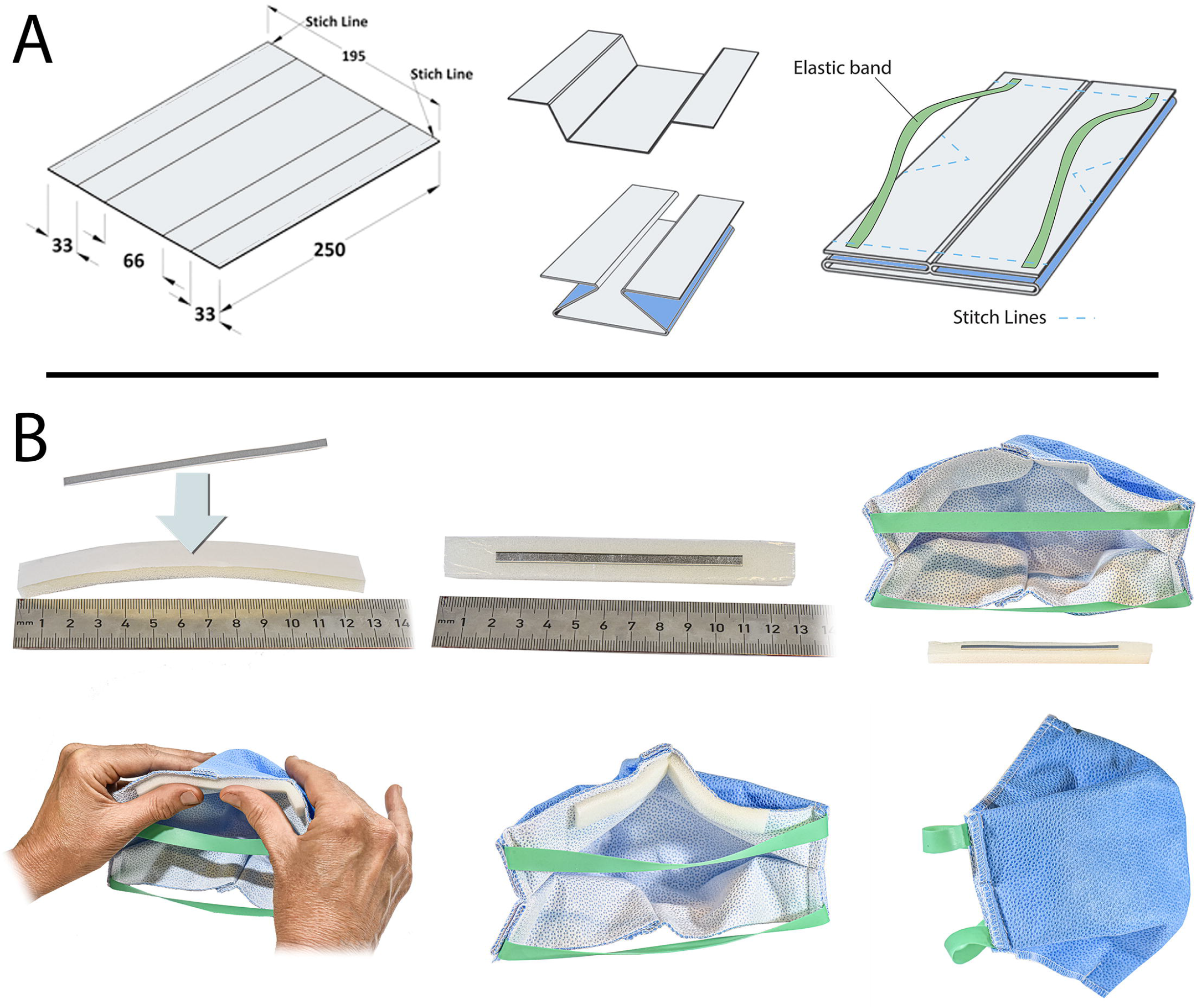
A possible design for fabricating a respirator mask with the sterile isolation material. (A) The layers of the Halyard sterile isolation material are attached by a stitch line on both longitudinal lengths of the sheets. The sheet is folded to provide alignment with the face, and thereby ensure the respirator mask has an adequate fit when worn. The elastics (Resistance Band, Matchu Sport BV) is laser-cut to a width of 13/32 inch (10mm), and a length of 7 7/8 inch (200mm) and attached at the inside of the respirator mask. A single stitch line at the bottom ensures that the surface of the respirator mask stays separated from the mouth and allows to adjust the size of the respirator mask for -and by- anybody. (B) For the nose clip a 0.5mm thick aluminum strips (Al 99.5%, 1050A) is used, cut to a length of 3 1/2 inch (90mm) and a width of 5/32 inch (4mm). A neoprene strip with adhesive is used to hold the noseclip in place, and adhered to the inner-top side of the respirator mask. The fit of the mask was tested using an FT-30 Fit Test from 3M.

In other studies, the suitability of alternative materials for use as respirator masks has been investigated. For instance, a regular tea cloth provides some protection, but remains a factor 50 below the FFP2 requirements [20]. A recent study analyzed the filtering efficiencies of various conventional fabrics to assess possible materials for the SARS-CoV-2 pandemic [21]. The results from this study contributed to understand the efficiencies for alternative materials for home-made masks and showed that widely available materials such as cotton and chiffon displayed filtering efficiencies above 95%, for larger, but not for smaller particles. Our study was specifically aimed to allow for solutions in hospitals and other critical environments and provide a solution in case of a limited supply of high quality protective respirator masks.

## Data Availability

The data referred to in the manuscript is summarized in Table 1 and in the supplementary data tables.

## Acknowledgements

The authors would like to thank the Development Department of the LUMC, Huybert van der Stadt (for photography), and André van der Zee (for assistance on measurements). We thank G.D. Block (UCLA), J.S. Takahashi (Southwestern University), and G.A. Fitzgerald (University of Pennsylvania) for providing general comments regarding the paper. We are particularly grateful to Nuri Cano (tailor) for pointing us in the right direction, and without whom this work had not been undertaken.

